# Effects of Side-Effect Risk Framing Strategies on COVID-19 Vaccine Intentions: A Randomized Controlled Trial

**DOI:** 10.1101/2021.10.12.21264877

**Authors:** Nikkil Sudharsanan, Caterina Favaretti, Violetta Hachaturyan, Till Bärnighausen, Alain Vandormael

**Author notes:** Corresponding author: Nikkil Sudharsanan, **Email:**.

## Abstract

Fear over side-effects is one of the main drivers of COVID-19 vaccine hesitancy. We conducted a pre-registered randomized controlled trial among 8998 individuals to examine the effects of different ways of framing and presenting vaccine side-effects on individuals’ willingness to get vaccinated. We found that adding a descriptive risk label (“very low risk”) next to the numerical side-effect and providing a comparison to motor vehicle mortality increased participants’ willingness to take the COVID-19 vaccine by 3.0 percentage points (p = 0.003) and 2.4 percentage points (p = 0.049), respectively. These effects were independent and additive and combining both framing strategies increased willingness to receive the vaccine by 6.1 percentage points (p < 0.001). Mechanistically, we find evidence that these framing effects operate by increasing individuals’ perceptions of how safe the vaccine is. Our results reveal that low-cost side-effect framing strategies can meaningfully affect vaccine intentions at a population level.

## Introduction

Vaccination is one of the main strategies for controlling the COVID-19 pandemic. However, vaccination rates have slowed and are far from target levels in countries like the United States and the United Kingdom. For example, in the United States, the share of the population that is fully vaccinated went from 1.8% in February 2021 to 40.4% in May 2021 but has since only risen to 54% in the following four months *(1)*. While not as stagnant, there is a similar pattern in the United Kingdom, where the share of the population that is fully vaccinated was just 65.6% as of September 27th, 2021 *(1)*. These vaccination trends are insufficient to prevent the spread of COVID-19, especially the Delta-variant, which has re-ignited the pandemic in both countries *(1–3)*.

Vaccine hesitancy is not the result of a single homogenous cause and can vary for different individuals and population groups. For example, recent studies have identified several potential reasons for COVID-19 vaccine hesitancy, including a low perceived risk of COVID-19 infection and concern around how quickly the vaccines were developed (*4–7*). A common finding across these studies is that fear and concern about vaccine side-effects is an important reason for vaccine hesitancy. These concerns were potentially heightened by widespread media coverage of vaccine side-effects in April and May 2021, along with the pausing of vaccination efforts in several countries due to this media coverage (*8, 9*). Although COVID-19 vaccine side-effects rates are extremely low (1 per 100,000 people vaccinated with AstraZeneca in the European Union) (*10*), these rates were often presented by the media without context (*9, 11–13*), likely leading some individuals to reject vaccine uptake (*14*). In addition, the pausing of global COVID-19 vaccination efforts may have sent a strong signal that side effects are a major cause of concern, thus increasing vaccine hesitancy.

Addressing public concerns over vaccine side-effects will be a key component of efforts to improve vaccine use in the United States, United Kingdom, and globally -- especially as new vaccines are released. There is a large body of evidence in the health communication and the behavioral sciences that shows that how risks are framed and presented to individuals can affect their perceptions of its severity and ultimately their behavior (*15–17*). For example, studies have shown that whether numerical risks are presented as percentages or natural frequencies (e.g. 1% compared to 1 out of 100), with a comparison to a commonly understood but different risk (e.g. the risk of motor-vehicle mortality), using infographics (e.g. visually showing the numbers of individuals in the numerator and denominator of a risk), with descriptive labels (e.g. putting “very low risk” or “high risk” next to the numerical risk) can all influence behavior. Therefore, simple changes to the framing of COVID-19 vaccine side-effect risks may have a meaningful influence on the number of people who choose to get vaccinated. Such “nudges” are viewed positively by policymakers since they are often affordable to implement and have the potential for wide-population reach (*18*).

These types of framing effects do not affect behavior by changing deeply held attitudes and beliefs, or by creating strong incentives for a certain behavior. Rather, research has shown that individuals have a limited cognitive ability to process and internalize risk information, especially rare risks like COVID-19 side-effects, and that individuals use mental guides or “heuristics” to make sense of risk information that ultimately guides their decisions (*19–21*). Therefore, framing effects work by modifying the heuristics that individuals use to understand a risk in a way that does not strongly affect their incentives or remove their agency.

What remains unknown to date, however, is what the effect of such framing effects are on COVID-19 vaccine intentions, which framing strategies are most promising, and whether such nudges can even influence COVID-19 vaccination behavior. Given that vaccination has become socially and politically charged, affecting hesitancy may no longer be amenable to light-touch nudge and framing interventions. Relatedly, it is an open question whether such framing effects will still be effective given that individuals have already experienced months of side-effect-related media coverage.

To answer these questions, we conducted a randomized controlled trial evaluating the effect of three framing strategies on COVID-19 vaccine intentions among 8988 adults ages 18+ (4502 adults in the United States and 4496 in the United Kingdom). We presented participants information on a hypothetical “future COVID-19 vaccine” including information on its side-effect rate (we chose a side-effect rate that was comparable to the current vaccines) (*22, 23*). We examined three side-effect framing strategies: what is the effect of adding a qualitative risk label next to the numerical risk, what is the effect of adding a comparison group (along with which comparison group is most effective), and for those with comparison groups, what is the effect of framing the comparison in relative rather than absolute terms (absolute comparisons of small risks may be cognitively harder for individuals to process than relative comparisons). Based on a pre-registered and published analysis plan (*24*), we then evaluated the effect of these framing strategies on two outcomes: self-reported willingness to take the hypothetical vaccine, and as a measure of the mechanism of our framing effects, individuals’ perceived safety of the vaccine.

## Results

### Effects of vaccine side-effect framing strategies on COVID-19 vaccine intentions

Figure 1 presents the results of the three main framing strategies on the proportion of participants that report that they would take the hypothetical vaccine (the results are presented on the percentage point scale). Among all the strategies, we find that adding a simple qualitative risk label (“very low risk”) next to the numerical risk increased vaccine intentions by 3.0 percentage points (95% CI: 1.0, 4.9; p = 0.003; control group mean: 65%). For our two comparison strategies, we found that adding a comparison to motor-vehicle mortality (Effect size: 2.4 percentage points; 95% CI: 0.001, 4.7; p = 0.049; control group mean: 65%) had an impact on vaccine intentions but no evidence of an effect of adding a comparison to COVID-19 mortality (Effect size: 0.8 percentage points; 95% CI: -1.6, 3.2; p = 0.496; control group mean: 65%). This is a surprising result as we expected the comparison to COVID-19 mortality to be more salient both because it is substantially higher than motor-vehicle mortality and because it is the form of mortality directly related to vaccination. In the US, for example, motor vehicle mortality in 2020 was 12 per 100,000 population while COVID-19 mortality in the same year was 170 per 100,000 population. Lastly, going against our expectation, we did not find evidence that relative, compared to absolute, framings of comparisons had a large impact on willingness to take the hypothetical future COVID-19 vaccine (Effect size: 1.3 percentage points; 95% CI: -1.1, 3.6; p = 0.285; control group mean: 67%).

**Figure 1.**
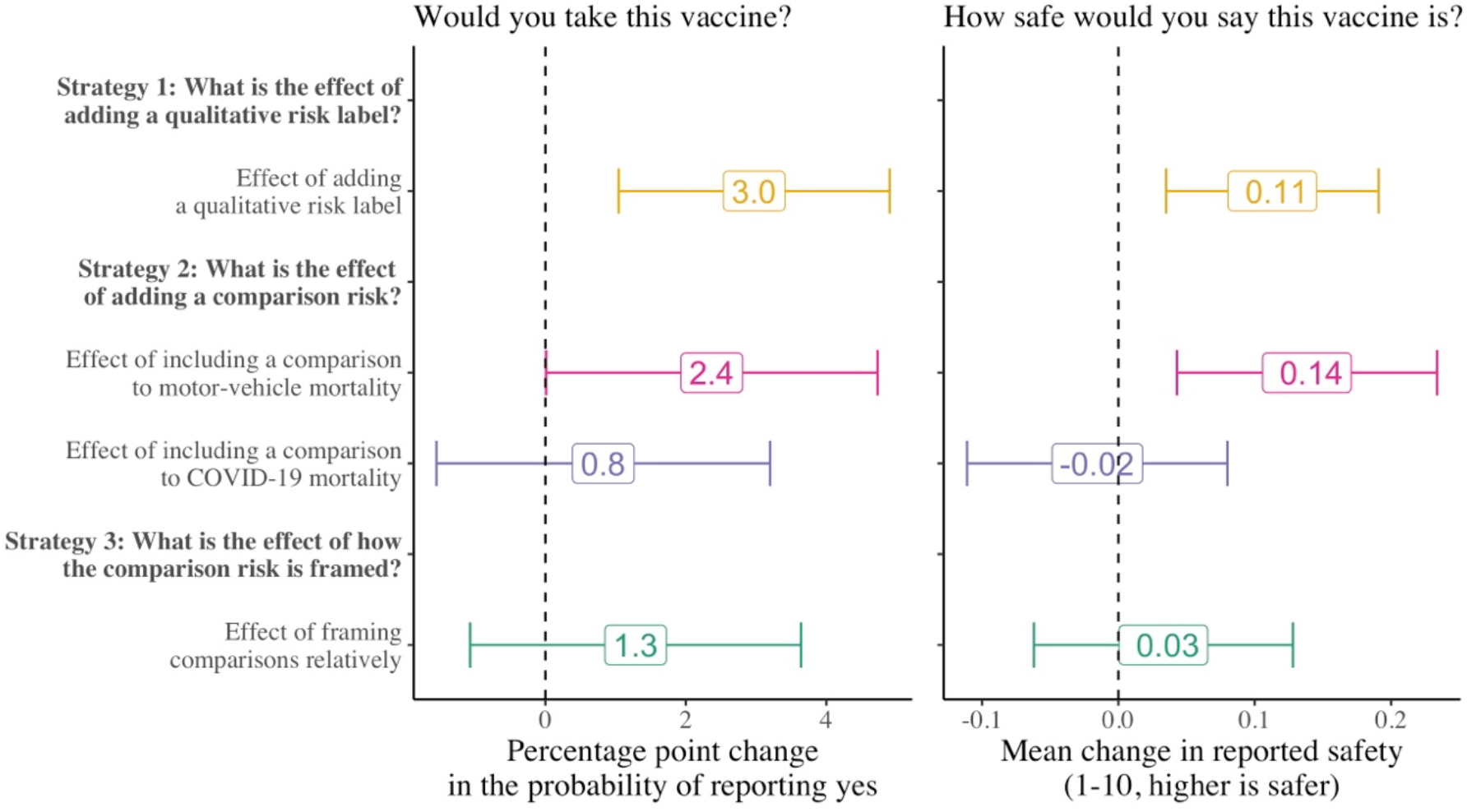
Effect of the vaccine side-effect framing strategies on the probability that participants report that they would take the hypothetical vaccine and their perceptions of how safe the hypothetical vaccine is (N = 8998 for Strategy 1 and 2); Strategy 3 is estimated only among those that received a comparison risk (N = 5998). Control means for strategies 1 and 2 are 65% and 67% for strategy 3.

Figure 1 also presents the effect of the strategies on participants’ perceptions of how safe the vaccine is (measured on a scale of 1-10). This secondary outcome serves to investigate whether the effects we observed on vaccine intentions were driven through perceptions of safety (providing more information may affect vaccine intentions, for example, by increasing individuals’ trust in the source of information rather than directly affecting their perceptions of vaccine safety). Although the magnitude of the effect sizes is small, we find that the pattern of the framing effects on perceptions of vaccine safety closely mirrors the primary effects on willingness to take the vaccine. This suggests that perceptions of safety are indeed one of the pathways through which these framing effects operate, although our analyses do not exclude the possibility that by providing more information, these framing strategies also affect vaccine intentions by increasing the perceived trustworthiness or reliability of the information.

### Are the two effects substitutes, additive, or synergistic?

An important question is how effective these strategies are when combined. For example, it could be the case that vaccine intentions are only movable by a fixed margin, such that even when both qualitative risk labels and comparison groups are used, the resulting change in vaccine intentions is less than the sum of the independent effects. Conversely, the effects may be synergistic such that combining strategies leads to larger effects than the sum of the independent effects. To assess this consideration, in a non-pre-registered analysis, we estimated the effect of adding both a qualitative risk label and comparison to motor vehicle mortality on willingness to take the hypothetical COVID-19 vaccine (Table 1). We find that at a minimum, the effects are independent and additive, with some indication that they may even be synergistic. Compared to those that received neither a quality risk label nor a comparison risk (N = 1499; control mean: 63%), those that received both strategies were 6.1 percentage points (95% CI: 2.8, 9.5; p < 0.001) more likely to state that they would take the vaccine.

**Table 1.** Independent and combined effects of risk labeling and motor-vehicle mortality comparisons on willingness to take a hypothetical COVID-19 vaccine.

|  | Effect Size<br>(percentage points) | p-value |
| --- | --- | --- |
| Effect of risk labeling<br>(ref: no risk label)<br>*N* = 8998 | 3.0 | 0.003 |
| Effect of motor-vehicle comparison<br>(ref: no comparison)<br>*N*=8998 | 2.4 | 0.049 |
| Effect of both risk labeling and a motor-vehicle<br>comparison<br>(ref: no risk label nor comparison)<br>*N*=3002 | 6.1 | <0.001 |

*Notes:* Outcome: "Would you take this vaccine?" (yes =1, others = 0). As per our pre-analysis plan, all regressions include covariates for age, sex, education, and country. Sample sizes for the relative to absolute comparison and effect of both labeling and a motor-vehicle mortality comparison are smaller since they are only estimated among subset of the total sample. P-values are from two-tailed t-tests.

### Differences by country, sex, and age group

Figure 2 examines whether the effects of these framing strategies vary by country, age, and sex. The coefficients in each figure show the interaction effect on the treatment with the main heterogeneity characteristic and are thus interpreted as how much larger is the framing effect for a particular group compared to another on the log-odds scale. There are several reasons to suspect that there may be differences in the magnitude of framing effects across groups. Contexts where vaccines have become highly politicized like the US may be less responsive to framing effects than in places like the UK. Older individuals may use different heuristics for assessing risk and thus respond differently to framing effects (although the direction of this is theoretically ambiguous); and on average, men and women may have different risk thresholds for what they deem to be risky or not (*25–27*), which may affect how the framing effects impact behavior.

**Figure 2.**
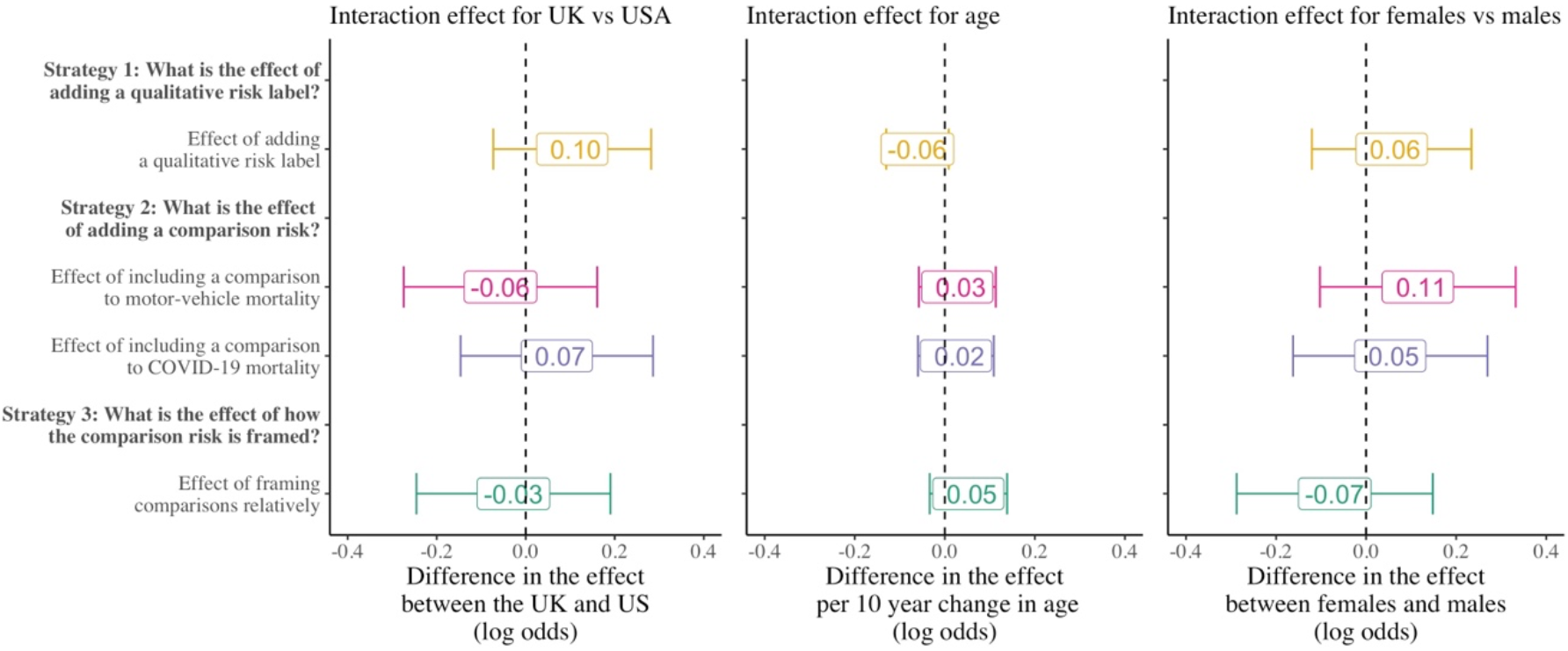
Differences in the framing effects by country, age, and sex. Coefficients are the interaction effect of the main heterogeneity characteristic with the indicator for each treatment strategy and are presented as coefficients on the log-odds scale from logistic regression models.

We do not find evidence that the framing strategies vary across country, age, or sex. Importantly, due to our sample sizes, we cannot rule out the possibility of potentially small interaction effects. Therefore, our results should be interpreted as not finding evidence for any large differences.

### Robustness

We find no evidence of imbalances in sociodemographic characteristics across the experimental groups (Supplemental Tables S1-S3) and our main results remain unchanged after excluding these characteristics as control variables in the main regression analyses (Supplemental Table S4). Our results are also consistent when we use linear probability rather than logistic regression models (Supplemental Table S4) and when we examine the outcome as a four-category ordinal (rather than binary) variable (Supplemental Table S5).

## Discussion

We found that adding a simple descriptive risk label (“very low risk”) next to the numerical side-effect increased participants’ willingness to take the COVID-19 vaccine by 3.0 percentage points (p = 0.003). Providing a comparison to motor vehicle mortality increased COVID-19 vaccine willingness by 2.4 percentage points (p = 0.049). Importantly, we found that these effects were independent and additive: participants that received both a qualitative risk label and comparison to motor-vehicle mortality were 6.1 percentage points (p < 0.001) more likely to report willingness to take a vaccine compared to those who did not receive a label or comparison. This is an important and meaningful effect at the population level, where even small changes in vaccination rates can have large health consequences (*28*). These results are especially reassuring considering the low cost and ease of implementing such framing strategies. Based on the effects on perceptions of vaccine safety, we find support for the hypothesis that these effects work by modifying individuals’ judgments about how safe the vaccine is. Taken together, our results reveal that despite increasingly strong vaccination hesitancy and exposure to large amounts of vaccine information, low-cost side-effect framing strategies can meaningfully affect vaccination intentions at a population level. Given that vaccination for COVID-19 will likely remain an important priority for the foreseeable future, these insights can be valuable for increasing the uptake of future vaccination efforts.

## Materials and Methods

### Study Approvals, Registration, and Pre-Analysis Plan

Prior to recruiting any participants, we received ethical approval for the study from the Medical Faculty of Heidelberg University Ethics Committee (#S-443/2021), registered the trial, including the outcomes and treatments, on the German Clinical Trials Registry (#DRKS00025551), and published a trial protocol with a pre-analysis plan (*24*). All the analyses and results we present here are in line with our pre-analysis plan.

The study here is missing one secondary sub-analysis that we registered as part of the protocol and pre-analysis plan. This additional analysis was intended to be based on a comparison of the main study participants (the 8998 individuals who form the current study) to an additional 3000 participants. The reason for the omission of this sub-analysis is that there was an error in the study text for these additional 3000 participants. This error, however, had no impact on the 8998 participants recruited for the main study presented here, and as stated in our pre-analysis plan, this omitted analysis was only intended as a supplementary analysis.

### Study Population, Eligibility, Inclusion, and Exclusion Criteria

This was an online-based randomized controlled trial study. To be enrolled in the trial, participants had to meet the following inclusion criteria be 18 years or older, have current residence in the United States or the United Kingdom, and be able to speak English. Participants who did not meet the inclusion criteria were not eligible to participate and were excluded from the study.

### Participant Recruitment

We used Prolific (*29*), an online-based service for recruiting participants for online-based research studies to recruit the study participants. Prolific participants are paid for their participation (we paid each participant £0.63/$0.88 for the expected 5-minute completion time). Within Prolific, we used a stratified quota sampling procedure to match the education distribution of our sample to the general population (stratified by sex and country) based on available data for each country (*30,31*). We aimed to recruit a total of 9000 total participants, split evenly by country and sex. 99% of our participants were recruited between 30 July 2021 and 10 August 2021. To reach our target quotas by sex and education, the final 1% of participants were recruited between 11 August 2021 and 4 October 2021.

### Description of the Experiment

We previously published the protocol for our trial with a description of the experiment (2*4)*. On the Prolific platform, potential participants were provided information that the aim of the study was to understand their willingness to take COVID-19 vaccines, the risks and benefits of the study, and how they could contact the researcher (and/or the human subjects review board at the Heidelberg University). After consenting on Prolific, participants were redirected to the Gorilla platform, an interactive web-based service for conducting behavioral science experiments. On the Gorilla page, we provided additional information on data protection. For participants that agreed to participate, we first collected basic sociodemographic information and then set up the experiment by telling participants that we are going to show them information on a hypothetical future COVID-19 vaccine and would like to know how willing they would be to take it. At this stage, we emphasized that their answers cannot be linked back to them in any way. Participants were then presented information on the vaccine and its side effect rate on a single page. The main experimental component was how the vaccine side effect rate was presented to participants.

### Randomization

We used a factorial randomization design to assign participants to three main factors. Factor 1 was whether there was a qualitative risk label saying “very low risk” next to the numerical risk or not. Factor 2 was whether the risk was presented with no comparison, a comparison to motor-vehicle mortality, or a comparison to COVID-19 mortality. Factor 3 was only among that received a comparison risk and varied whether the comparison was presented in an absolute or relative way. We randomized participants to each factor independently (stratified by country), such that, for example, which comparison group a participant received did not depend on whether they received a risk label or not. This means that the proportion of participants that received a risk label will be balanced across the treatment groups for the comparison risks. We repeated this independent randomization procedure for whether the comparison risk is presented as an absolute or relative comparison among the 2/3rds of the sample that were randomized to receive any comparison risk. Similarly, this means that conditional on receiving any comparison risk (either motor vehicle or COVID-19 mortality), the proportion of participants that received each type of comparison, and the proportion that received a labeled risk will be balanced across the treatment groups for absolute and relative framings. Note that in practice, the Gorilla algorithm handled the randomization using a stratified randomization approach to prevent chance imbalances in the joint distribution of the factors.

### Outcomes

Our main outcome was a binary variable for whether participants reported “yes” to the question of “Would you take this vaccine if it were made available to you?” Importantly we asked participants to answer as if they had not been vaccinated even if they already received some form of vaccination. In the Supplemental Table S5, we show the main study results using a four-category ordinal response rather than a binary response variable and find no change to our conclusions. Our secondary outcome was participants’ perception of how safe the vaccine is, based on a scale of 1 (extremely unsafe) to 10 (extremely safe). This secondary outcome served to determine if the effects we saw on vaccine willingness were at least partly driven through perceptions and judgments of the vaccine’s safety.

## Data Availability

The datasets generated during and/or analysed during the current study and the analysis codes used to produce the figures and tables are available in the Open Science Framework repository, https://doi.org/10.17605/OSF.IO/HQNKR.

## Acknowledgments

None.

## Author Contributions

NS wrote the manuscript. NS and AV designed the trial and developed the questionnaires. NS, AV, CF, and VH carried out the experiment. NS performed the analysis. All authors read and approved the final manuscript.

