## Supplemental Information for "Effects of Side-Effect Risk Framing Strategies on COVID-19 Vaccine Intentions: A Randomized Controlled Trial"

### Supplementary Information

#### Materials and Methods

##### *Missing Data*

We had complete data for all 4502 participants from the USA (the extra two participants above our target of 4500 was the result of how Prolific and Gorilla recruit individuals). 4 individuals from the UK sample did not complete the experiments and were therefore excluded due to missing data. This resulted in a final sample of 8998 individuals.

##### *Statistical Analyses*

We first assessed if the randomization was successful by comparing means and proportions of the main sociodemographic variables across the experimental arms for each of the randomization factors.

Next, as per our pre-analysis plan, we estimated the main effects of the risk label and comparison group strategies using the following logistic regression model:

$$ln\left(\frac{\theta}{1-\theta}\right) = \alpha_0 + (\alpha_1 * CM_i) + (\alpha_2 * MVM_i) + (\alpha_3 * QL_i) + \boldsymbol{\zeta_i} \ (1)$$

In this model, θ is the probability of reporting "Yes" to take the hypothetical vaccine, $CM_i$ is a binary indicator for whether participant *i* was shown a comparison to COVID-19 mortality, $MVM_i$ is a binary indicator for whether participant *i* was shown a comparison to motor-vehicle mortality, $QL_i$ is a binary indicator for whether participant *i* was shown a qualitative risk label, and $\boldsymbol{\zeta_i}$ is a vector of covariates, including age, sex, education, and country. Our main effects of interest are $\alpha_1$, $\alpha_2$, and $\alpha_3$ respectively.

We then estimated the effect of a relative compared to absolute comparison framing by estimating the following regression just among those that received either the motor-vehicle or COVID-19 mortality comparison:

$$ln\left(\frac{\theta}{1-\theta}\right) = \beta_0 + (\beta_1 * Rel_i) + \boldsymbol{\zeta}_i(2)$$

Here $Rel_i$ is a binary indicator for receiving the relative framing and $\beta_1$ is the corresponding effect of this framing on vaccine intentions compared to an absolute comparison.

Based on our pre-registered analyses, we found evidence that adding a qualitative risk label and adding a comparison to motor-vehicle mortality both increased the likelihood that participants reported that they would take the vaccine. In a non-pre-registered analysis, we then assessed whether these two independent effects were additive by estimating the effect of receiving both strategies relative to receiving neither.

To determine whether the main effects varied by country, sex, and age, we re-estimated these two main regressions including interaction terms between the main experimental indicators and the heterogeneity variables (separately for each heterogeneity variable).

Lastly, to determine whether these effects were partly mediated by perceptions of vaccine safety, we estimated regressions (1) and (2) using a continuous indicator for reported safety as the main outcome variable. As robustness and sensitivity analyses, we re-estimated each regression without controls for age, sex, country, and education, using an ordinal logistic regression model with a four-category outcome variable for vaccine intention rather than a binary indicator for "yes," and using linear probability rather than logistic regression models.

To ease the interpretation, we present coefficients as average marginal effects for all logistic regressions (as per our pre-analysis plan).

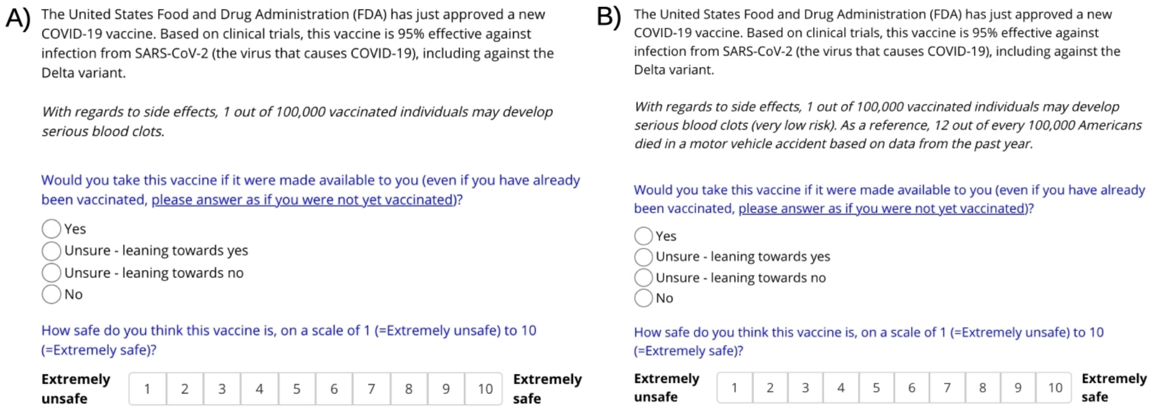


**Fig. S1.** Screenshots of the online, Gorilla-based, experiment content for 2 of the 10 possible experimental combinations (A) no risk label, no control group; (B) risk label, comparison to motor-vehicle mortality, absolute comparison.

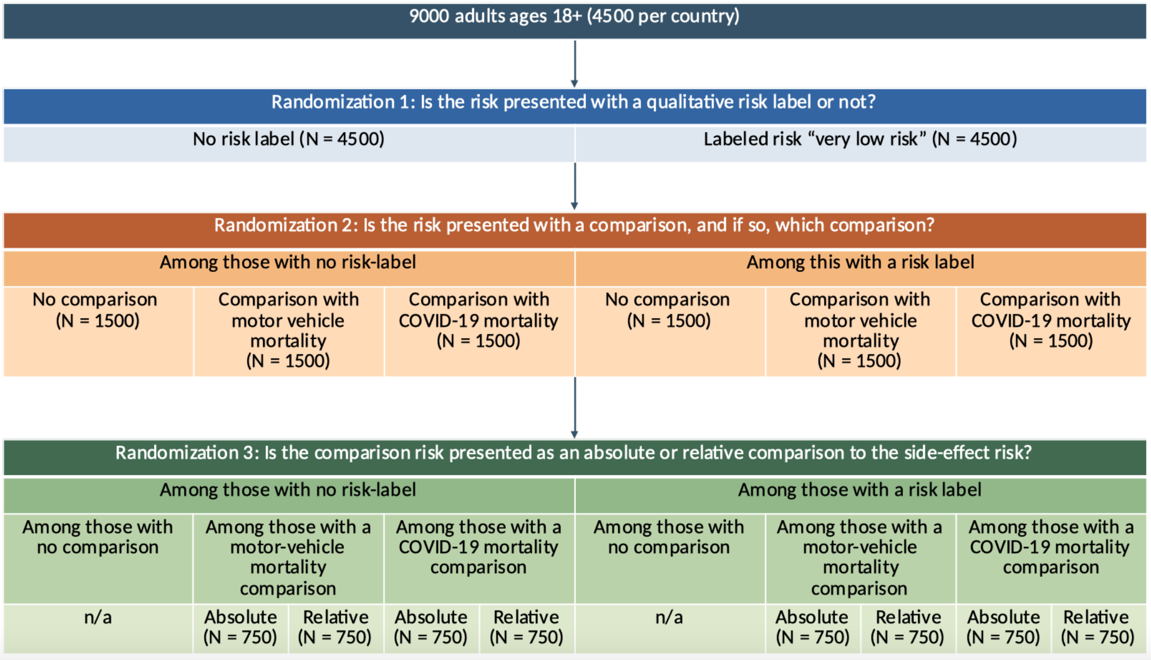


**Fig. S2.** Detailed randomization diagram across the main study factors. We have shown the randomization here with sample sizes referring to both countries combined; in practice, we conducted this procedure stratified by country such that each cell in the diagram has exactly 50% USA and 50% UK observations.

**Arm 1**

The United States Food and Drug Administration (FDA) has just approved a new COVID-19 vaccine. Based on clinical trials, this vaccine is 95% effective against infection from SARS-CoV-2 (the virus that causes COVID-19), including against the Delta variant.

*With regards to side effects, 1 out of 100,000 vaccinated individuals may develop serious blood clots.*

**Arm 2**

The United States Food and Drug Administration (FDA) has just approved a new COVID-19 vaccine. Based on clinical trials, this vaccine is 95% effective against infection from SARS-CoV-2 (the virus that causes COVID-19), including against the Delta variant.

*With regards to side effects, 1 out of 100,000 vaccinated individuals may develop serious blood clots. As a reference, 12 out of every 100,000 Americans died in a motor vehicle accident based on data from the past year.*

**Arm 3**

The United States Food and Drug Administration (FDA) has just approved a new COVID-19 vaccine. Based on clinical trials, this vaccine is 95% effective against infection from SARS-CoV-2 (the virus that causes COVID-19), including against the Delta variant.

*With regards to side effects, 1 out of 100,000 vaccinated individuals may develop serious blood clots. As a reference, this is 1/12th of the risk of dying in a motor vehicle accident based on data from the past year.*

**Arm 4**

The United States Food and Drug Administration (FDA) has just approved a new COVID-19 vaccine. Based on clinical trials, this vaccine is 95% effective against infection from SARS-CoV-2 (the virus that causes COVID-19), including against the Delta variant.

*With regards to side effects, 1 out of 100,000 vaccinated individuals may develop serious blood clots. As a reference, 170 out of every 100,000 unvaccinated Americans died of COVID-19 based on data from the past year.*

**Arm 5**

The United States Food and Drug Administration (FDA) has just approved a new COVID-19 vaccine. Based on clinical trials, this vaccine is 95% effective against infection from SARS-CoV-2 (the virus that causes COVID-19), including against the Delta variant.

*With regards to side effects, 1 out of 100,000 vaccinated individuals may develop serious blood clots. As a reference, this is 1/170th of the risk of COVID-19 mortality among unvaccinated Americans based on data from the past year.*

**Arm 6**

The United States Food and Drug Administration (FDA) has just approved a new COVID-19 vaccine. Based on clinical trials, this vaccine is 95% effective against infection from SARS-CoV-2 (the virus that causes COVID-19), including against the Delta variant.

*With regards to side effects, 1 out of 100,000 vaccinated individuals may develop serious blood clots (very low risk).*

**Arm 7**

The United States Food and Drug Administration (FDA) has just approved a new COVID-19 vaccine. Based on clinical trials, this vaccine is 95% effective against infection from SARS-CoV-2 (the virus that causes COVID-19), including against the Delta variant.

*With regards to side effects, 1 out of 100,000 vaccinated individuals may develop serious blood clots (very low risk). As a reference, 12 out of every 100,000 Americans died in a motor vehicle accident based on data from the past year.*

**Arm 8**

The United States Food and Drug Administration (FDA) has just approved a new COVID-19 vaccine. Based on clinical trials, this vaccine is 95% effective against infection from SARS-CoV-2 (the virus that causes COVID-19), including against the Delta variant.

*With regards to side effects, 1 out of 100,000 vaccinated individuals may develop serious blood clots (very low risk). As a reference, this is 1/12th of the risk of dying in a motor vehicle accident based on data from the past year.*

**Arm 9**

The United States Food and Drug Administration (FDA) has just approved a new COVID-19 vaccine. Based on clinical trials, this vaccine is 95% effective against infection from SARS-CoV-2 (the virus that causes COVID-19), including against the Delta variant.

*With regards to side effects, 1 out of 100,000 vaccinated individuals may develop serious blood clots (very low risk). As a reference, 170 out of every 100,000 unvaccinated Americans died of COVID-19 based on data from the past year.*

**Arm 10**

The United States Food and Drug Administration (FDA) has just approved a new COVID-19 vaccine. Based on clinical trials, this vaccine is 95% effective against infection from SARS-CoV-2 (the virus that causes COVID-19), including against the Delta variant.

*With regards to side effects, 1 out of 100,000 vaccinated individuals may develop serious blood clots (very low risk). As a reference, this is 1/170th of the risk of COVID-19 mortality among unvaccinated Americans based on data from the past year.*

**Fig. S3.** Screenshots of each experimental arm for United States participants. Note that we have labeled the arms for the figure, but participants were not shown this label. Reference mortality information for motor-vehicle and COVID-19 fatalities were taken from the Centers for Disease Control and Prevention *(2,3)*.

**Arm 1**

The Medicines and Healthcare products Regulatory Agency (MHRA) has just approved a new COVID-19 vaccine. Based on clinical trials, this vaccine is 95% effective against infection from SARS-CoV-2 (the virus that causes COVID-19), including against the Delta variant.

*With regards to side effects, 1 out of 100,000 vaccinated individuals may develop serious blood clots.*

**Arm 2**

The Medicines and Healthcare products Regulatory Agency (MHRA) has just approved a new COVID-19 vaccine. Based on clinical trials, this vaccine is 95% effective against infection from SARS-CoV-2 (the virus that causes COVID-19), including against the Delta variant.

*With regards to side effects, 1 out of 100,000 vaccinated individuals may develop serious blood clots. As a reference, 2.6 out of every 100,000 individuals in the UK died in a motor vehicle accident based on data from the past year.*

**Arm 3**

The Medicines and Healthcare products Regulatory Agency (MHRA) has just approved a new COVID-19 vaccine. Based on clinical trials, this vaccine is 95% effective against infection from SARS-CoV-2 (the virus that causes COVID-19), including against the Delta variant.

*With regards to side effects, 1 out of 100,000 vaccinated individuals may develop serious blood clots. As a reference, this is almost 1/4th of the risk of dying in a motor vehicle accident based on data from the past year.*

**Arm 4**

The Medicines and Healthcare products Regulatory Agency (MHRA) has just approved a new COVID-19 vaccine. Based on clinical trials, this vaccine is 95% effective against infection from SARS-CoV-2 (the virus that causes COVID-19), including against the Delta variant.

*With regards to side effects, 1 out of 100,000 vaccinated individuals may develop serious blood clots. As a reference, 108 out of every 100,000 unvaccinated individuals in the UK died of COVID-19 based on data from the past year.*

**Arm 5**

The Medicines and Healthcare products Regulatory Agency (MHRA) has just approved a new COVID-19 vaccine. Based on clinical trials, this vaccine is 95% effective against infection from SARS-CoV-2 (the virus that causes COVID-19), including against the Delta variant.

*With regards to side effects, 1 out of 100,000 vaccinated individuals may develop serious blood clots. As a reference, this is 1/108th of the risk of COVID-19 mortality among unvaccinated individuals in the UK based on data from the past year.*

**Arm 6**

The Medicines and Healthcare products Regulatory Agency (MHRA) has just approved a new COVID-19 vaccine. Based on clinical trials, this vaccine is 95% effective against infection from SARS-CoV-2 (the virus that causes COVID-19), including against the Delta variant.

*With regards to side effects, 1 out of 100,000 vaccinated individuals may develop serious blood clots (very low risk).*

**Arm 7**

The Medicines and Healthcare products Regulatory Agency (MHRA) has just approved a new COVID-19 vaccine. Based on clinical trials, this vaccine is 95% effective against infection from SARS-CoV-2 (the virus that causes COVID-19), including against the Delta variant.

*With regards to side effects, 1 out of 100,000 vaccinated individuals may develop serious blood clots (very low risk). As a reference, 2.6 out of every 100,000 individuals in the UK died in a motor vehicle accident based on data from the past year.*

**Arm 8**

The Medicines and Healthcare products Regulatory Agency (MHRA) has just approved a new COVID-19 vaccine. Based on clinical trials, this vaccine is 95% effective against infection from SARS-CoV-2 (the virus that causes COVID-19), including against the Delta variant.

*With regards to side effects, 1 out of 100,000 vaccinated individuals may develop serious blood clots (very low risk). As a reference, this is nearly 1/4th of the risk of dying in a motor vehicle accident based on data from the past year.*

**Arm 9**

The Medicines and Healthcare products Regulatory Agency (MHRA) has just approved a new COVID-19 vaccine. Based on clinical trials, this vaccine is 95% effective against infection from SARS-CoV-2 (the virus that causes COVID-19), including against the Delta variant.

*With regards to side effects, 1 out of 100,000 vaccinated individuals may develop serious blood clots (very low risk). As a reference, 108 out of every 100,000 unvaccinated individuals in the UK died of COVID-19 based on data from the past year.*

**Arm 10**

The Medicines and Healthcare products Regulatory Agency (MHRA) has just approved a new COVID-19 vaccine. Based on clinical trials, this vaccine is 95% effective against infection from SARS-CoV-2 (the virus that causes COVID-19), including against the Delta variant.

*With regards to side effects, 1 out of 100,000 vaccinated individuals may develop serious blood clots (very low risk). As a reference, this is 1/108th of the risk of COVID-19 mortality among unvaccinated individuals in the UK based on data from the past year.*

**Fig. S4.** Screenshots of each experimental arm for United Kingdom participants. Note that we have labeled the arms for the figure, but participants were not shown this label. Reference mortality information for motor-vehicle fatalities were taken from the Department of Transportation *(4)* and COVID-19 from the Public Health England (*5*).

**Table S1.** Balance table of baseline participant characteristics between the No Risk Label and Risk Label treatment arms.

| | No Risk Label (N=4496) | Risk Label (N=4502) | P-value |
|---|---|---|---|
| Female (n, %) | 2224 (49.5%) | 2197 (48.8%) | 0.542 |
| Age (mean years, SD) | 31.7 (12.6) | 31.6 (12.7) | 0.623 |
| Education (n, %) | | | 0.836 |
| Less than secondary education | 320 (7.1%) | 319 (7.1%) | |
| Completed secondary education | 1097 (24.4%) | 1107 (24.6%) | |
| College | 1392 (31.0%) | 1409 (31.3%) | |
| Bachelor's degree | 1143 (25.4%) | 1100 (24.4%) | |
| More than bachelor's degree | 544 (12.1%) | 567 (12.6%) | |

*Notes:* P-values for Gender and Education correspond to a chi-squared test and a two-sided t-test for Age.

**Table S2.** Balance table of baseline participant characteristics between the No comparison, Comparison with motor-vehicle mortality, and Comparison with COVID-19 mortality treatment arms.

| | No Comparison (N=3000) | Comparison with motor vehicle mortality (N=3002) | Comparison with COVID-19 mortality (N=2996) | P-value |
|---|---|---|---|---|
| Female (n, %) | 1478 (49.3%) | 1452 (48.4%) | 1491 (49.8%) | 0.547 |
| Age (mean years, SD) | 31.8 (12.8) | 31.4 (12.5) | 31.6 (12.7) | 0.516 |
| Education (n, %) | | | | 0.873 |
| Less than secondary education | 210 (7.0%) | 202 (6.7%) | 227 (7.6%) | |
| Completed secondary education | 717 (24.0%) | 744 (24.8%) | 743 (24.8%) | |
| College | 931 (31.0%) | 933 (31.1%) | 937 (31.2%) | |
| Bachelor's degree | 769 (25.6%) | 753 (25.1%) | 721 (24.1%) | |
| More than bachelor's degree | 373 (12.4%) | 370 (12.3%) | 368 (12.3%) | |

*Note*. P-values for Gender and Education correspond to a chi-square test and an ANOVA-test for Age.

**Table S3.** Balance table of baseline participant characteristics between the Absolute Risk and Relative Risk treatment arms.

| | Absolute Risk (N=2997) | Relative Risk (N=3001) | P-value |
|---|---|---|---|
| Female (n, %) | 1473 (49.1%) | 1470 (49.0%) | 0.918 |
| Age (mean years, SD) | 31.4 (12.4) | 31.6 (12.8) | 0.543 |
| Education (n, %) | | | 0.897 |
| Less than secondary education | 222 (7.4%) | 207 (6.9%) | |
| Completed secondary education | 737 (24.6%) | 750 (25.0%) | |
| College | 934 (31.2%) | 936 (31.2%) | |
| Bachelor's degree | 743 (24.8%) | 731 (24.3%) | |
| More than bachelor's degree | 361 (12.0%) | 377 (12.6%) | |

*Note*. P-values for Gender and Education correspond to a chi-square test and a two-sided t-test for Age.

**Table S4.** Robustness of main paper results to alternative regression types and the inclusion/exclusion of covariates

| | Logistic regression models with results presented as average marginal effects | | Linear probability models | |
|---|---|---|---|---|
| | With covariates (main paper results) | Without covariates | With covariates | Without covariates |
| Effect of labeled risk compared to unlabeled risk | 3.0 pp ($p = 0.003$) | 3.0 pp ($p = 0.003$) | 3.0 pp ($p = 0.003$) | 3.0 pp ($p = 0.003$) |
| Effect of comparison to motor-vehicle mortality compared to no comparison | 2.4 pp ($p = 0.049$) | 2.4 pp ($p = 0.051$) | 2.4 pp ($p = 0.0498$) | 2.4 pp ($p = 0.052$) |
| Effect of comparison to COVID-19 mortality compared to no comparison | 0.8 pp ($p = 0.496$) | 0.7 pp ($p = 0.568$) | 0.8 pp ($p = 0.497$) | 0.7 pp ($p = 0.571$) |
| Effect of relative comparison compared to absolute comparison | 1.3 pp ($p = 0.285$) | 1.3 pp ($p = 0.277$) | 1.3 pp ($p = 0.286$) | 1.3 pp ($p = 0.277$) |
| Effect of both risk labeling and a motor-vehicle comparison compared to no labeling or comparison | 6.1 pp ($p < 0.001$) | 6.1 pp ($p < 0.001$) | 6.1 pp ($p < 0.001$) | 6.1 pp ($p < 0.001$) |

*Notes*: Outcome: "Would you take this vaccine?" (yes =1, others = 0). As per our pre-analysis plan, covariates include age, sex, education, and country. Sample sizes for the relative to absolute comparison (N = 5998) and effect of both labeling and a motor-vehicle mortality comparison (N = 3002) are smaller since they are only estimated among subset of the total sample.

**Table S5.** Main paper results estimated using ordinal logistic regression models with a four-category outcome specification rather than logistic regression models with a binary outcome

| | Odds Ratio (p-value) |
|---|---|
| Effect of labeled risk compared to unlabeled risk | 1.15 (p = 0.002) |
| Effect of comparison to motor-vehicle mortality compared to no comparison | 1.11 (p = 0.050) |
| Effect of comparison to COVID-19 mortality compared to no comparison | 1.03 (p = 0.541) |
| Effect of relative comparison compared to absolute comparison | 1.05 (p = 0.327) |
| Effect of both risk labeling and a motor-vehicle comparison compared to no labeling or comparison | 1.31 (p < 0.001) |

*Note.* Outcome: "Would you take this vaccine?" (No, Unsure - leaning towards no, Unsure - leaning towards yes, Yes). As per the main paper results and pre-analysis plan, all regressions include controls for age, sex, country, and education. Sample sizes for the relative to absolute comparison (N = 5998) and effect of both labeling and a motor-vehicle mortality comparison (N = 3002) are smaller since they are only estimated among subset of the total sample.
